# Learning about COVID-19 across borders: Public health information and adherence among international travellers to the UK

**DOI:** 10.1101/2021.08.04.21261333

**Authors:** Shenghan Cai, Tingting Zhang, Charlotte Robin, Clare Sawyer, Wendy Rice, Louise E. Smith, Richard Amlôt, G. James Rubin, Lucy Yardley, Matthew Hickman, Isabel Oliver, Helen Lambert

**Author notes:** Corresponding author: Professor Helen Lambert. Population Health Sciences, Bristol Medical School, University of Bristol, Bristol, UK.

## Abstract

**Objective:** Public health control measures at borders have long been central to national strategies for the prevention and containment of infectious diseases. Travel was inevitably associated with the rapid global transmission of COVID-19. In the UK, public health authorities took action to reduce risks of travel-associated spread by providing public health information at ports of entry. This study aims to understand individual risk assessment processes, decision making, and adherence to official advice among international travellers; to provide evidence to inform future policy on the presentation of public health information to facilitate safer international travel.

**Study design:** This study is a qualitative study evaluation.

**Method:** Semi-structured interviews were conducted to investigate risk assessment processes, decision making, and adherence to official Public Health England (PHE) advice among travellers.

**Results:** Participants regarded official advice as adequate at the time, despite observing differences between the intervention measures implemented in the countries of departure. Participants however also described adopting precautionary measures including self-isolation and the use of face coverings that went beyond official advice, and variability in the extent to which they adhered to guidance on contacting health authorities. Adherence to official guidance was informed by the perceived salience of specific transmission possibilities and containment measures assessed in relation to participants’ social and institutional environments.

**Conclusion:** Analysis of travellers’ reported motivations demonstrates that responses to public health advice constitute a proactive process of risk assessment and rationalised decision-making that incorporates consideration of living situation, trust in information sources, correspondence with cultural logics, and willingness to accept potential risk to self and significant others in guiding preventive action. Our findings concerning international passengers’ understanding of, and compliance with, official advice and mitigation measures provide valuable evidence to inform future policy and we provide recommendations on the presentation of public health information to facilitate safer international travel. Access to a central source of regularly updated official information would help minimise confusion between different national guidelines. Greater attention to the differentiated information needs of diverse groups in creating future public-facing guidance would help to minimise the uncertainties generated by receipt of generic information.

## Introduction

The significant transmission risks associated with travel mean that public health disease control measures at borders have long been central to national strategies for the prevention and containment of infectious diseases. Since early 2020 COVID-19 cases have surged across successive countries and continents^1^ despite country-specific attempts at containment though widely varying travel restrictions. With over a third of the world’s population having been placed under lockdown restrictions, the socioeconomic impact of COVID-19 may be causing the largest global recession in history^2^, with decades of progress and development at risk, especially for low- and middle-income countries^3^.

Although interventions such as mass vaccination may ultimately achieve the aim of containing the global spread of COVID-19, public health information and guidance on behavioural measures remains vital, particularly when national health systems have faced increasing pressure and new variants with increased transmissibility are creating threats to global containment and effective vaccination^4,5^. Knowledge and information can support the public to adopt measures that will mitigate or prevent transmission and to date, these behavioural non-pharmaceutical interventions are still the key measures to control the spread of COVID-19 across the world.

As the first cases of COVID-19 were reported, public health authorities in the UK took action to reduce the risk of travel-associated spread through provision of public health advice at ports of entry starting on 23^rd^ January 2020. On 25^th^ January Public Health England (PHE) activated the Airport Public Health Monitoring Operations Centre to monitor all direct flights from China to London Heathrow Airport (LHR), and on 29^th^ January operations were extended to include all international direct flights to London (Heathrow and Gatwick) and Manchester. Measures directed at passengers travelling from affected countries into England included (see supplementary document):

- a broadcast message to passengers made on incoming aircraft, to encourage travellers to report relevant symptoms
- posters containing COVID-19 related public health advice displayed at Arrivals in these airports
- leaflets containing the same advice provided to passengers by airlines on board flights and/or made available on arrival.

These measures remained in place until extensive travel restrictions were implemented on 23^rd^ March as part of a national lockdown - with UK residents prohibited from travelling abroad unless they had a permitted reason to do so, while returning travellers were required to quarantine for 14 days.

As the UK and other countries’ vaccination programmes progress and lockdown measures in certain countries are eased, future provision for travelling across borders has again become a pressing concern. Amid discussion of vaccine passports and pressure from the travel industry and national economies dependent on tourism to ease restrictions, passengers will be left navigating the differing rules, guidance, and social norms between countries. Historical evidence on international passengers’ views on, and compliance with, official advice and mitigation measures can provide valuable evidence to inform policy on the presentation of public health information to facilitate safer international travel. Our previous evaluation of the official advice on COVID-19 provided to international travellers reported on the impact and effectiveness of these communication materials in the early stage of the pandemic^6^. Drawing on qualitative data collected prior to the implementation of travel restrictions, this paper presents findings on individual assessment of risk, decision making, and adherence practices among travellers arriving into the UK by air who were confronted with potentially unfamiliar public health information. These findings provide wider insights into interactions between official advice and individual behaviour and indicate possible improvements to public health guidance for future international travel.

## Methods

A mixed-methods study was conducted to evaluate the public health information provided to international travellers arriving into the UK. Passengers aged 18 years and over arriving at LHR airport on three scheduled flights from China and Singapore in March 2020 were recruited for a cross-sectional survey and semi-structured interviews regarding their experience and understanding of the official guidance they received, as well as their subsequent actions.

The survey (n=121) collected information regarding knowledge of COVID-19 symptoms and help-seeking behaviours, views on public health advice provided by PHE, and participants’ demographic information^6^. Respondents indicated willingness to be interviewed by recording their contact details on the questionnaire and consenting to participate when contacted on arrival; 15 of these respondents subsequently participated in semi-structured interviews.

One-to-one semi-structured telephone interviews were conducted in April 2020 in either English or Mandarin, according to participant preference. All interviews were audio-recorded with consent. Two topic guides were used; one which explored participants’ experience and views regarding COVID-19 information received during travel was used with all participants, while the second explored experiences relating to self-isolation if a participant reported having self-isolated following either potential exposure or development of COVID-19 symptoms.

Key information from each interview was summarised by researchers in field notes either during or immediately after the completion of interviews. Prior to data analysis, audio-recorded English interviews were transcribed verbatim and audio-recorded Mandarin interviews were transcribed directly into English.

### Data analysis

Interview transcripts were initially coded independently (by TZ, SC, CS, WR) using open coding, followed by collaborative development of an initial coding framework that was then used to index each transcript in NVivo 12 Pro. Codes that represented similar concepts were assembled into conceptual categories and themes. Common categories emerging across the transcripts indicated that all themes reached saturation.

## Results

Participants’ age range was 20 to 80 years; five were male and 10 were female. Among the 15 participants, 10 were permanent residents in the UK, five were visitors or temporary residents. Six participants were retired; five worked full-time; three were full-time students and one was unemployed. Most participants were British; all three Chinese participants spoke Mandarin and English and had read the official advice materials in both languages, while other participants only knew English and accordingly, only read the English version of the PHE materials. Six participants have a degree in higher education. One participant reported symptoms of COVID-19 after arrival.

### Perceptions of public health measures

14 participants recalled obtaining the official information from PHE on COVID-19 (leaflets and/or posters) in flight or at the airport, while one participant reported only receiving local information at the port of departure. Most participants stated that they considered the UK response of providing official advice to be adequate at the time. However, they also reported finding the situation at the UK airport on arrival to be dramatically different from the official public health intervention measures they had experienced at their place of departure (Table 1, quote 1).

**Table 1.** Quotes of international traveller’s perceptions of public health measures.

| Number | Quotes |
| --- | --- |
| 1 | At China's airport... you need to fill in a Health Declaration Card... they will check your temperature; all the [airport] staff were fully equipped with PPE... [in the UK] only when I went through the customs that I saw the hand sanitiser there. They [airport staff] didn't wear face masks... So, in China, the official advice is wearing face masks and washing hands as often as possible. In the UK, as no one was wearing a face mask, it gave you the impression that things were not bad here. |
| 2 | At Heathrow, it was like nothing was wrong in the UK, so I think that causes a false sense of security, maybe if there was more of a presence, like information, temperature check, personnel etc, people might take it more seriously. I think a lot of people probably didn't take it seriously at the time. |
| 3 | They could have had thermal imaging cameras, they could have had medical staff in protective clothing there to talk to people whose temperature came up as above the norm, they could have then asked people in those conditions, if they met those conditions to isolate them. |
| 4 | When we came back through in March the taxi driver said that there hadn't really been too many more cases. So, at the time, how silly, it didn't seem to be as serious as we all now know it is. |
| 5 | We'd kind of already sort of lived with it, listened to it out there probably all through February [on the television], we were a bit ahead of the game if you like in terms of that we'd already seen it. |
| 6 | I think we found out enough ourselves and heard enough and talked about it between ourselves and came to the decisions that we did, so yeah, so I don't really think that there could have been any more advice at that time that would have made any difference because as I say that advice if you like that was coming out later on we were already doing because of being where we'd been I suppose. |
| 7 | Even if you self-isolate I think nobody quite realised how contagious this virus is, so when it means self-isolate it means self-isolate, it means don't touch anyone, you know, wear a mask. I think at the time little was really known about the virus. If you're going to redesign the leaflet again it might not only say to self-isolate but wear a mask, do not come into contact with each other, self-isolate but also practice social distancing as well to make sure that you do not give the virus to anybody else. |
| 8 | It was difficult because we'd had no idea what was going on anyway, because at that point it definitely felt a bit over the top, we didn't know how many cases were in the UK and you don't know if you've actually contracted it or not. I do know people that have come back from Italy, I think the advice then was you need to isolate yourself for a week but they just kind of took it as oh well I'll stay in my house, but I'll still be around my housemates and my family and stuff like that, so I think they need to actually take it seriously and just get used to not doing anything, being ok with not doing anything. |
| 9 | I can't think why you would not follow the official advice, at the time the number of people who had died from Coronavirus was rising and the numbers were unclear, but they were talking about one to two percent of the people who got infected may die. |

Summarised based on qualitative research data.

Based on their experiences on arrival, participants described complex understandings about the public health advice and measures they were exposed to. Even though COVID-19 related posters and leaflet stands had been set up at LHR, 12 participants had no recollection of seeing such posters or leaflets at all. Direct observation by our research team verified that the visibility of these information sources was very limited due to print size, colour, and placement^6^. Two participants recalled seeing hand sanitisers placed at the airport. Participants emphasised the amount of information and measures being reported in the media or at the airport in their departure country, as well as the protection measures that were applied on board the flight, in contrast with the situation at the arrival port. British participant also expressed frustration and concern that airport staff at the disembarkation point had not provided detailed instructions regarding COVID-19 situation in the UK and possible protection measures.

Most participants described their uncertainty about the COVID-19 situation in the UK at the time, as there was limited evidence of active intervention to contain transmission associated with travel on their arrival. An airport without visible containment measures was considered by the participants as signalling good containment of the virus in the UK; nevertheless, participants also described their need for reassurance in the absence of detailed instructions and protection measures (Table 1, quotes 2-4).

All participants were eager to acquire information regarding the pandemic and official advice about how to protect themselves and their family. Participants reported that they were proactively searching for advice and information on traditional and social media to understand the changing situation at ports of departure and arrival, evaluate potential risks, and identify measures they should take for international travel (Table 1, quotes 5 & 6).

Based on their experience in the country of departure, some participants also pointed out that following official advice was voluntary in the UK and noted that ‘advice and rules imposed by the British government are already very loose’. Participants also stated their awareness of the vacuum of scientific knowledge and detailed guidance at the early stage of the pandemic, which provided a space for interpretation of official advice regarding actions people should take (Table 1, quotes 7-9).

### Precautionary measures

Although participants described the official advice as adequate, based on their experience from more severe outbreaks overseas, some participants took actions beyond those recommended in official advice to reduce the risk of infection and transmission. When arriving in the UK, some participants voluntarily self-isolated or tried to maintain social distance by skipping social activities and gatherings as a precaution, which were not required according to the UK official advice. Participants expressed their concerns over the seemingly ‘business as usual’ situation in the UK, which contradicted their experience in places with established outbreaks, and chose to take extra precautions such as staying indoors, socially distancing, wearing masks, and monitoring their temperatures daily, even though they were not symptomatic (Table 2, quotes 1 & 2).

**Table 2.** Quotes of international traveller’s perceptions of precautionary measures.

| Number | Quotes |
| --- | --- |
| 1 | We sort of self-isolate anyway when we came back. Nobody told us to do it. Because of the precautions we had taken and because we hadn't really hardly been with anybody; the chances of us giving others anything were miniscule. |
| 2 | I felt scared when I came back here. Because everyone in China took it seriously, but no one took it seriously here in the UK. When I go out, I wear a face mask, a pair of goggles and a pair of disposable gloves. I cover myself tight. |
| 3 | We wouldn't have put anybody at risk if we really thought that were was a chance, but we just didn't want it on us. We knew the chances were absolutely miniscule, but we took the decision that we wouldn't see anybody, so that was family and friends, for over a week after we got back. |
| 4 | There was a kind of stigma with Singapore and South Korea and a few other Southeast Asian countries. We wanted to make everybody aware that if they did get something it possibly wouldn't have come from us because we'd been self-isolating. |
| 5 | I think they [official advice] were only saying like contact 111 basically if your symptoms get worse but obviously, they're inundated, but certainly had we got symptoms on those first few days after we got back then that's what we would have done because as I say at that time it was still quite a new thing here. |
| 6 | Someone told me it [NHS 111] is constantly engaged. I would have hoped to know how to contact the NHS effectively in the case that I was infected. Maybe I could have been given a few more telephone numbers? This kind of information enabling me to have access to medical treatment would have given me a sense of security. |
| 7 | We're luckier than most and if I want to go down and take a walk, I sort of can. I think if somebody is locked up in a one-bedroom flat in London it will be horrible. |
| 8 | I think providing they have sufficient support in their communities there is no reason at all why anybody should not self-isolate. |
| 9 | I don't think so [received any official support]. But the school sent us emails. It offered a backup option. If we encountered any problem, we could contact the school in the case of need. The school made us feel that they are quite protective. |
| 10 | The quarantine officer [at Heathrow airport] told me: don't worry, it is ok, don't be nervous; it is no use to wear a face mask. Social distancing was not taken at that time. I felt there would be loopholes and hidden risk. The outbreak has worsened in the UK, I think it has something to do with the measures taken then. [The UK] Airports were free zones at that time, hidden risk reveals itself when you entirely depend on voluntary. |
| 11 | We've seen [on the TV] about the mask doing more harm than good so we had to keep saying no, we're not going to get one because it's going to do more harm than good. |
| 12 | There wasn't any clear evidence to say an ordinary mask would prevent you picking up germs and if you did pick up a germ it would multiply inside the mask. So even though we had masks in our bags, but we decided we'd use the hand gel, but we didn't want to wear the masks. |
| 13 | The concern is, are you depriving the NHS of masks, when you're buying them for yourself when there is such a short global supply. So, there's a difficulty and a dilemma in just do I wear a mask. There is also a flipside to wearing masks, if you happen to have the mask and if your mask happens to pick up droplets from somebody else, your mask might become contagious. |

Summarised based on qualitative research data.

Several participants also mentioned their motivation for these voluntary precautionary behaviours as being in part related to potential stigma associated with the possibility of being seen as ‘contagious’ due to being travellers who had just returned from epidemic hotspots. They demonstrated willingness to follow the official containment measures even though they were not required to do so, both to avoid such stigma and to protect their family and friends (Table 2, quotes 3 & 4).

Study participants were asked whether they knew what they should do if they had developed symptoms or visited pandemic hotspots; and whether they were familiar with the NHS 111 service. All participants mentioned difficulties accessing the service, while participants were visitors or temporary residents in the UK were more concerned about the vagueness of advice itself and their uncertainty about whether formal support was available for non-citizens (Table 2, quotes 5 & 6).

Alongside calling NHS 111, British participants noted the additional options of contacting their GP or seeking support from their local communities or officials. They viewed it as easy to access any resources they might need to follow the official advice, such as support from their local authority or the community and were appreciative of this. However, interviewees who were visitors or temporary residents (such as foreign citizens travelling on business or studying in the UK) reported relying on personal or social networks, social media, and employers, as well as NHS 999 in case of emergency, but stated that they had limited contact with and support from local authorities and communities (Table 2, quotes 7-9).

Participants also demonstrated contrast in responses and selection of which official advice they should follow, especially regarding the use of face coverings. Some participants emphasised their concerns that airport staff did not wear face coverings and thought this was due to cultural and policy differences from their departure countries. They had already adopted face coverings as a daily prevention and protection measure, and strongly suggested that wearing masks should be included in official UK guidance (Table 2, quote 10).

Conversely, some participants noted they did not believe in the value of using face coverings due to official announcements from countries such as New Zealand and Singapore, on the lack of clear evidence of their effectiveness. Others shared an ambiguous ‘wait-and-see’ attitude towards face coverings, while a few strongly believed that people should use them to control the spread of the virus (Table 2, quotes 11-13).

## Discussion

Clear and actionable official information can help to shape the public’s understanding of the pandemic and promote public adherence to official advice^6,7,8^. The COVID-19 pandemic has been characterised by rapid changes to official guidance and advice, in response to newly emerging scientific evidence and shifting local situations^9^. Early in the pandemic, COVID-19 knowledge, information, and discourse was largely bounded by the national situation within each country and constrained by the limited scientific knowledge about the SARS-CoV-2 virus^10^. The transnational experience of the international travellers in this study exposed them to multiple versions of official information and interventions to contain transmission that became sources both of valued knowledge and of uncertainty, or confusion. This was exacerbated at this early stage of the pandemic by frequent changes in public health guidance and implementation of control measures.

This study has demonstrated that at the beginning of the pandemic in the UK, international travellers were relatively confident about their knowledge and action on self-protection against the virus and proactively used information acquired from multiple international sources to minimise their risk of exposure and transmission - despite limited detail in formal and official advice on arrival. However, it is also clear that there were differences in logic behind individual interpretations and precautionary actions, especially in the usage of face coverings, linked to discrepancies in public health information and socio-cultural norms between countries of origin and arrival.

In taking additional precautions beyond those officially advised by UK authorities at the time, many of the international travellers in our study were not following local official advice even at this early stage in the pandemic, albeit because they were ‘ahead of the game’. Arriving from countries that had already experienced severe outbreaks, these travellers had acquired knowledge from the places where travel originated and adopted additional precautionary measures that went beyond local (UK) recommendations. Since the COVID-19 situation was unclear in the UK at that time, arrival ports had not instituted control measures, leading many arriving passengers to question the seriousness of the UK government’s response to the pandemic^6^. The explicit public health information they received was less comprehensive than, and in some areas contradicted, official responses in the countries where travel originated. One consequence of these cross-national discrepancies in public health guidance and control measures was that international travellers were left to rely on their own judgement to navigate the salience and appropriateness of the differing rules, guidance, and social norms across national borders.

Our findings show that participants responded to the need to assimilate and interpret sometimes inconsistent information from multiple sources in response to spatial variations and changing pandemic situations across national borders, by developing their own ‘customised’ information set of public health guidance to inform action. Their responses to official UK public health advice constituted a proactive risk assessment and rationalised decision-making process whereby their living situation in UK, trust in information source, correspondence with cultural logics and degree of willingness to accept potential risk to self and significant others all contributed to choosing the most appropriate advice to guide preventive action. The international travellers who took actions that went beyond the extent of official advice in the UK at the beginning of the pandemic; even though these actions can be regarded as not strictly adherent to UK government advice at the time, they constituted effective precautionary measures. Although study participants repeatedly described the basis of their actions while manoeuvring across borders as “common sense”, their interpretations of multiple versions of official information and consequent behaviour are clearly context-based and consistent with their sociocultural capital.

These tailored responses are informed but not entirely determined by population-level determinants of vulnerability, such as reproduction rate of virus or prevalence within specific population groups and geographical regions. Thus, participants with a British background reported being relatively confident in following the UK official advice at the time, while participants who were visitors or temporary residents reported greater uncertainty and were more likely to maintain transmission-minimising precautions by adopting different sets of protective actions such as mask-wearing and non-mandated self-isolation. Most of these participants indicated that the element of official guidance on ‘calling NHS 111’, in the absence of detailed instruction or explanation of the advised action, had caused confusion and uncertainty. This was particularly clear among young participants who were visitors or temporary residents without pre-existing health issues and lacked knowledge about health services in the host country. They also reported lacking connections with local communities and being highly reliant on other members of their own community for support. Accordingly, these participants had concerns about the vagueness of official advice and expressed preference to adopt precautionary measures drawing on prior knowledge derived from their countries of origin and with sociocultural resources, rather than with officially designated processes in the UK. Although there were no significant differences in participants’ awareness and knowledge about COVID-19 symptoms and self-protection measures^6^, compared with other travellers, participants who were visitors or temporary residents had willingly adopted additional preventive measures that were not required according to UK official guidance at the time, to minimise their risk of exposure and transmission, and maintained such caution after their arrival. Such measures might have helped to reduce infection and transmission risks; however, longer stay visitors may have faced other adverse impacts as the pandemic continued due to their limited integration into local structures for practical, social, and emotional support, as well as difficulties in accessing health care. These difficulties which heighten potential vulnerability may also apply to resident minority communities in the UK and especially to more recent migrants. Studies have shown higher rates of COVID-19 exposure among minority communities due to socioeconomic disparities^11,12^, and individuals of these communities have faced more barriers on adhering to official advice during self-isolation and national lockdowns^8,13^.

Several limitations exist for this study. First, our sample of international travellers was limited due to the rapid implementation of travel restrictions and border closures in multiple countries in response to the rapidly changing situation of COVID-19 pandemic. In addition, all participants of telephone interviews reported that they had essential business or reasonable needs to travel abroad, our findings therefore may not be generalisable to those travelling for leisure purposes. However, the study benefits from being an early evaluation of adherence behaviour and responses to public health guidance among international travellers in the pandemic. Most importantly, it provides an opportunity to learn from the opening stages of the pandemic about how people navigate between differing national rules and guidance across borders. It provides insights that can inform recommendations for improving information provision and hence individual adherence for public health benefit.

### Public health implication

While international travellers were conscious of the potential risks associated with travel and keen to mitigate those risks, it is important to ensure that international travellers get information and official guidance on the key restrictions and preventive measures for the country to which they are travelling. The provision of a centralised and regularly updated official information hub would help to minimise possible confusion between different sources of guidance. In addition, incorporating into public health guidance, an explicit recognition and explanation of why international travel inevitably entails exposure to official public health containment measures, regulations, and sources of public health knowledge that differ across national jurisdictions may, in itself, help to reassure travellers.

Our findings suggest that in addition to differences in official information and regulations on COVID-19 across national borders, factors including information consistency, sociocultural norms, perceived risks and benefits, and availability of support from both official and unofficial sources all affect people’s adherence to official public health advice. In line with Denford *et al*.^8^, our findings indicate that individual adherence to official public health advice involves a decision-making process to select the health threats and measures to contain them that are considered particularly salient in the social context and institutional environment within which people are living. The process of selecting appropriate measures to adopt requires adaptation of non-specific official advice from different sources and self-management. Greater attention to the differentiated information requirements of diverse groups of international travellers in the design of future public health guidance – for example, through the provision of tailored information for dual residents, short-stay business and leisure travellers, and long-stay migrant workers and students - would help to minimise the uncertainties generated by receipt of generic advice which does not readily fit with individual circumstances. It is likely that these categories of travellers could be anticipated in preparation for future cross-border outbreaks, and key aspects of generic guidance pre-written to facilitate the rapid generation of tailored guidance in response. Clarification of financial and other support measures available in the destination country for both short- and long-stay travellers would enhance adherence to requirements, such as mandatory quarantine periods supported by testing, when these are not institutionally provided but depend on voluntary action.

## Data Availability

The data generated or analysed during this study are included in this article; data are available from authors from Public Health England or University of Bristol upon reasonable request.

## Disclosure statement

No potential conflict of interest was reported by the author(s).

## Declarations

This study was a form of service evaluation; therefore, no ethics committee approval was required. This was confirmed by the PHE Research Ethics and Governance Group.

All participants were informed about the purpose of this study and their participation was voluntary. The participants were told they could withdraw at any time without giving any reasons or facing any consequences. They agreed to the interview being recorded and that any identifiable information would be removed. Consent was obtained from participants to use anonymous quotes to be published. Signed and verbal consent were obtained at the beginning of both survey and interviews.

## Acknowledgements

We wish to thank all participants who contributed to this study and all staff in London Heathrow Airport for their kind support. The authors acknowledge support from the NIHR Health Protection Research Unit in Behavioural Science and Evaluation at University of Bristol, in partnership with Public Health England (PHE). L.Y. is a NIHR Senior Investigator and partly supported by NIHR Applied Research Collaboration (ARC)-West, NIHR Health Protection Research Unit (HPRU) in Behavioural Science and Evaluation, and the NIHR Southampton Biomedical Research Centre (BRC). M.H., I.O. and H.L. are supported by the NIHR Health Protection Research Unit (HPRU) in Behavioural Science and Evaluation at the University of Bristol in partnership with Public Health England. L.S. and J.R. are supported by the NIHR HPRU in Emergency Preparedness and Response at King’s College London in partnership with Public Health England. C.R. is affiliated to the National Institute for Health Research Health Protection Research Unit (NIHR HPRU) in Emerging and Zoonotic Infections at the University of Liverpool in partnership with Public Health England (PHE), in collaboration with the Liverpool School of Tropical Medicine and the University of Oxford, the NIHR HPRU in Gastrointestinal Infections at the University of Liverpool in partnership with PHE, in collaboration with the University of Warwick and the NIHR HPRU in Behavioural Science and Evaluation at the University of Bristol, in partnership with PHE.

## Funding

This study was funded by NIHR on behalf of the Department of Health and Social Care. The views expressed are those of the authors and not necessarily those of the NIHR, the Department of Health and Social Care, or PHE. The views expressed are those of the author(s) and not necessarily those of the NHS, the NIHR, the Department of Health or Public Health England.

## Notes

### Competing Interest Statement

The authors have declared no competing interest.

### Funding Statement

This study was funded by NIHR on behalf of the Department of Health and Social Care. The authors acknowledge support from the NIHR Health Protection Research Unit in Behavioural Science and Evaluation at University of Bristol, in partnership with Public Health England (PHE). The views expressed are those of the authors and not necessarily those of the NIHR, the Department of Health and Social Care, or PHE.

